# C No More: A prospective single-arm study to evaluate the effectiveness of a nurse and peer-led mobile model of hepatitis C testing and treatment at community corrections offices

**DOI:** 10.1101/2025.10.05.25337369

**Authors:** Samara Griffin, Timothy Papaluca, Jacinta A Holmes, Bridget Reid, Anne Craigie, Jane Dicka, Sione Crawford, Amanda Callus, Mark Belzer, Tim Spelman, Margaret Hellard, Shelley Walker, Mark Stoové, Alexander J Thompson, Rebecca J Winter

## Abstract

**Background:** The community corrections population in Australia shares similar risk factors for hepatitis C virus (HCV) infection with people incarcerated in prisons, but without access to prison-based testing and treatment. While hepatitis C testing and treatment programs are well established in prison settings in Australia, little attention has been paid to equivalent programs in community corrections settings. C No More is a study to evaluate the acceptability and efficacy of a novel, mobile, nurse and peer-led model of hepatitis C testing and treatment at community corrections offices in Melbourne, Australia.

**Methods:** A clinically equipped van staffed by a hepatitis clinical nurse consultant and peer workers will spend scheduled periods parked adjacent to four community corrections offices in metropolitan Melbourne. People attending community corrections offices will be opportunistically approached by a peer worker and invited to undertake hepatitis C testing. Other individuals in the vicinity of the community corrections office may also be invited to access the service. Following enrolment, study staff will conduct hepatitis C point-of-care testing and clinical assessments in the van. Point-of-care HCV antibody tests will be used for initial screening, and where positive, point-of-care HCV RNA tests performed. Participants with self-reported HCV antibody will be reflexed to RNA testing. RNA positive participants will be assessed for rapid treatment initiation, and prescribed DAA treatment. Treatment dispensation will occur from the van or through a community pharmacy. Participants will be monitored throughout treatment and offered point-of-care HCV RNA testing four or more weeks after treatment completion to confirm sustained virological response (SVR-4+). The primary outcomes of this study are the proportion of those who are HCV RNA positive who initiate DAA treatment and the acceptability of this model of care.

**Discussion:** This study will provide evidence of acceptability, feasibility, and clinical effectiveness of a nurse and peer-led, mobile model of hepatitis C care at community corrections offices. The outcomes of this study will inform other models of care aiming to provide hepatitis C testing and treatment to individuals involved in the criminal justice system.

**Clinical Trial Registration:** This study is registered with the Australian New Zealand Clinical Trials Registry (ACTRN12623001043628). Date of registration: 27/09/2023.

## Background and rationale

In Australia, hepatitis C virus (HCV) is predominantly transmitted through the sharing or reusing of injecting drug equipment by people who inject drugs (1). People who inject drugs typically face multiple barriers accessing traditional modes of healthcare, including competing daily priorities, experiences of stigma and discrimination, and complicated health systems (2, 3).

The development of direct-acting antivirals (DAAs), a highly effective and well-tolerated treatment for hepatitis C, led the World Health Organization (WHO) to set goals to eliminate hepatitis C as a public health threat by 2030 (4). Despite widespread access to DAAs and strong early progress, substantial declines in treatment initiation threaten Australia’s progress to elimination (5). To achieve WHO elimination goals, rates of hepatitis C testing and treatment initiation must increase, particularly in groups who have not yet been reached with previous engagement strategies (5, 6). To reach these populations, adaptable, mobile, and patient-centred models of care are required in settings which people at risk of hepatitis C frequent (7).

Due to the criminalisation of drug use, people who inject drugs experience disproportionate rates of incarceration. As such, Australian prisons have an estimated hepatitis C prevalence of 8%, compared to 1% in the community (8), making prisons a key setting to engage people living with hepatitis C in care. Prison-based hepatitis C treatment programs in Australia have been highly feasible, clinically effective and cost-effective, providing care and treatment to a large number of people (9-13) . Prisons are increasingly important in progressing Australia’s hepatitis C elimination goals, with prison-based treatments accounting for 35% of hepatitis C treatments nationally in 2023, and declining at a lesser rate than community treatments (14, 15).

While the criminal justice system provides an opportunity to screen large numbers of people at-risk of hepatitis C, not everyone engaged in the criminal justice system is incarcerated. In Australia, 60% of the people in the corrections system are supervised in the community, including community corrections orders (probation) and parole (16). This group of people, whilst likely having similar risk profiles to people in prison, may never have had access to prison-based care. Further, some individuals who are incarcerated decline prison-based testing or have inadequate periods of incarceration to undergo testing, commence treatment, or arrange appropriate continuity of care on release. Individuals re-entering the community from prison are met with significant and immediate competing financial and social priorities; siloed prison and community healthcare systems further complicate linkage to hepatitis C care in the community by placing the onus on individuals to seek post-release hepatitis C care (13, 17-19).

There is considerable overlap of the various groups engaged in the corrections system. People may be released from prison on parole or serve combined prison and community supervision orders, while people on community corrections orders (with no prison component) can also be incarcerated due to breaching court-ordered conditions. Despite those on community corrections orders outnumbering those that are incarcerated (20) and significant overlap with the incarcerated population, there are currently few programs in Australia, and none in Victoria (Australia’s second most populous state) offering hepatitis C care in community corrections settings (21). In contrast to prison hepatitis programs, there is very little literature describing models of care for hepatitis C in community corrections populations internationally (22-25). There is great potential in providing hepatitis C care in community corrections settings, due to the population likely being at high risk of infection, however there is limited evidence about the impact, feasibility, and effectiveness of these services (13, 23, 26). Specifically, the best model of care in the community corrections setting is not clear; the contribution of factors such as location, testing and treatment strategies, and staffing of the service is unknown.

Hepatitis C care has traditionally required multiple blood tests and clinical appointments to diagnose active hepatitis C infection, for treatment assessment, to initiate DAA treatment, and then to confirm cure, contributing to extremely high attrition rates throughout this process (27). Government funded hepatitis C testing is current venepuncture based and requires an initial HCV antibody test and then an HCV RNA test for confirmation of current infection. Point-of-care testing can help address attrition by facilitating rapid same-day hepatitis C diagnosis at the site of clinical care. Point-of-care hepatitis C antibody and RNA testing provide results within an hour using capillary specimens, negating the need for venepuncture. Fingerstick blood sampling for testing is highly acceptable to people who inject drugs, as venepuncture requirements can be a barrier due to injecting related vein damage and venous access difficulties (28). Point-of-care hepatitis C tests can simplify models of care and increase engagement in care and linkage to treatment (29), allowing for non-clinically trained staff to conduct testing (29-31).

New approaches to providing hepatitis C care including community-based settings, and point-of-care testing have been shown to be effective, however these may not be as effective in the community corrections setting, with a population that may have had limited interaction with health services and may not trust health services. Involving peers in the design and delivery of hepatitis C services is critical for an appropriate healthcare model, as peer-led models of care have been demonstrated to increase engagement and trust and reduce feelings of judgement (32-35). Nurse-led models of care have an established role in hepatitis C treatment, with strengths including accessibility, cost-effectiveness and person-centred care (11, 36-38). As such, the flexibility of point-of-care testing is well suited for use by these important stakeholders, encouraging task-sharing and creating possibilities for broadening of hepatitis C models of care.

For Australia to reach hepatitis C elimination, new and adapted models of care that reach populations missed with current strategies is vital (14). The establishment of an effective hepatitis C care program to service the community corrections population therefore represents a key opportunity to expand hepatitis C testing and treatment for this population with unmet needs.

We hypothesise that a mobile outreach peer and nurse-led model of hepatitis C care at community corrections offices, utilising point-of-care testing and rapid pathways to treatment initiation will be clinically effective and acceptable.

### Objectives

The objectives of the C NO More study are to determine the:

1. Clinical effectiveness and feasibility of point-of-care hepatitis C testing and treatment at community corrections offices;
2. The acceptability of the model of care for participants.

### Study design

This is a single arm clinical trial to evaluate the effectiveness and feasibility of mobile rapid point-of-care hepatitis C testing and treatment initiation at community corrections offices. A medically equipped mobile van staffed by a hepatitis clinical nurse consultant and peer worker will attend selected community corrections sites. Participants will be recruited opportunistically at four designated community corrections sites over an 18-month period.

## Methods

### Participants, interventions, outcomes

This protocol follows the SPIRIT recommendations and checklist (39) (Supplementary 1).

### Study Setting

The study will be conducted in a mobile clinically equipped van, parked adjacent to four selected community corrections offices.

Community corrections offices manage and supervise individuals serving community-based sentences, including individuals on probation, parole, drug treatment orders, and community corrections orders. Supervision requirements and reporting regularity vary depending on the sentence and the individual. Requirements can include electronic tracking, urine monitoring for alcohol and other drug use, community-based work hours, and individual or group counselling. Individuals may be required to report daily, weekly, or monthly to corrections offices.

Community corrections offices are typically located centrally to social services and public infrastructure, including welfare and housing offices and public transport. In Victoria, there are 54 community corrections offices, 20 of which are in the Melbourne metropolitan area (40). As of June 2024, there were 9,650 individuals serving community corrections orders in Victoria (20). Four sites were selected within the Melbourne metropolitan area in Victoria, Australia as they were identified as having high target client throughput. The four selected sites are in the North, South-East, and West of the Melbourne metropolitan area.

### Eligibility Criteria

Whilst focusing on community corrections locations, recruitment at sites is not restricted to individuals under community supervision at the time of enrolment; members of the community within the vicinity of the mobile van can be invited to participate in the study.

### Study inclusion criteria

Aged ≥ 18 years;

Consenting to fingerstick testing for hepatitis C antibody and/or hepatitis C RNA;

Consenting to provide full name, date of birth, contact details; and

Has a Medicare number (Medicare is Australia’s publicly funded universal healthcare insurance scheme; a Medicare number is required to access publicly funded DAA treatment in Australia).

### Study exclusion criteria

Already engaged in treatment for hepatitis C infection.

Participants may be excluded if there is contraindication or caution to hepatitis C DAA treatment initiation, including:

- Receiving medications which are contraindicated to be co-administered with DAAs or has significant drug-drug interactions (DDIs);
- Pregnant or breastfeeding at the time of commencing DAA therapy; and
- Individuals with decompensated cirrhosis, prior treatment failure or prior significant adverse events to DAAs.

These individuals will be referred to the supervising tertiary hospital, St Vincent’s Hospital Melbourne, for specialist review. In addition, patients with suspected significant liver disease will also be referred to St Vincent’s Hospital for review.

### Diagnostic testing

#### Point-of-care hepatitis C testing

Point-of-care hepatitis C testing kits were provided by the National Point of Care Testing Program (41) including both antibody and RNA tests. Briefly, the national program supports point-of-care hepatitis C testing and includes access to consumables, as well as providing operator training, quality assurance programs, and support.

##### Point-of-care HCV antibody testing

Point of care HCV antibody testing will be performed by a trained clinical nurse consultant, study staff, or peer worker using the INSTI® HCV antibody test, or the Abbott Bioline® test. These tests are performed using a fingerstick collection of 50μL of capillary blood with a minivette. The INSTI® and Bioline® HCV antibody tests are manual, visually read, flow-through qualitative immunoassay tests, and provide a qualitative antibody result within 60 seconds (INSTI®) or 20 minutes (Bioline®). The INSTI® has 100% sensitivity and 99.7% specificity, and the Bioline® has 98.8% sensitivity and 100% specificity, both comparable with the performance of conventional laboratory tests (42, 43).

The INSTI® HCV antibody test received regulatory approval as a diagnostic test in 2024. As of July 2024, the Abbott Bioline® HCV antibody test had not been TGA approved; therefore, antibody testing will be conducted under clinical trial conditions (ACTRN12623001043628). Participants who self-report previous hepatitis C infection or prior DAA treatment will proceed directly to point-of-care HCV RNA testing.

##### Point-of-care HCV RNA testing

Point of care hepatitis C RNA testing will be performed by a trained clinical nurse consultant, study staff, or peer worker using the Cepheid GeneXpert ® HCV VL Fingerstick assay. This test is performed using a fingerstick collection of 100μL of capillary blood with a minivette and provides a quantitative result with a lower limit of detection of 100IU/mL for most HCV genotypes, with a result available within 60 minutes of sample collection. Participants who are hepatitis C RNA positive will undergo further assessment for treatment eligibility.

### Health assessments

Participants who test positive for hepatitis C RNA will be offered venepuncture for confirmatory HCV RNA and genotype testing, screening for other blood borne viruses (hepatitis B and HIV) and pre-treatment blood tests for non-invasive assessment of liver fibrosis. Participants who test positive for hepatitis B and/or HIV infection will be provided with post-test counselling by trained study staff and will be referred to appropriate health services for assessment. To reduce the burden of repetitive testing, the results of any available recent blood tests may be used. If all pathology tests are not able to be completed, this will be documented in the clinical data collection tool. Participants will not be excluded from treatment initiation if these tests cannot be performed.

### Liver fibrosis assessment

Participants who are hepatitis C RNA positive and undertake venepuncture will undergo liver fibrosis assessment. Initial fibrosis assessment will be performed using the aspartate transaminase to platelet ratio (APRI) score. If an individual has an APRI score >1.0, they will be referred for further evaluation with transient elastography (FibroScan®). Participants who do not undertake venepuncture, and have no recent pathology results available to calculate APRI score will be referred for a FibroScan® to assess baseline liver fibrosis, however clinical assessment of liver fibrosis will be conducted and the absence of a FibroScan® or APRI will not preclude treatment initiation.

### Hepatitis C treatment

Participants who test positive for hepatitis C RNA will be prescribed DAA treatment by the study team: the nurse consultant will undertake initial assessment and remotely consult with either a nurse practitioner or gastroenterologist for prescribing. Some participants with decompensated cirrhosis, complex DDIs or multiple prior DAA treatment failures will be referred for specialist gastroenterology review for DAA treatment initiation. DAA treatment will be prescribed per the Australian hepatitis C guidelines (44) (Figure 1).

**Figure 1.**
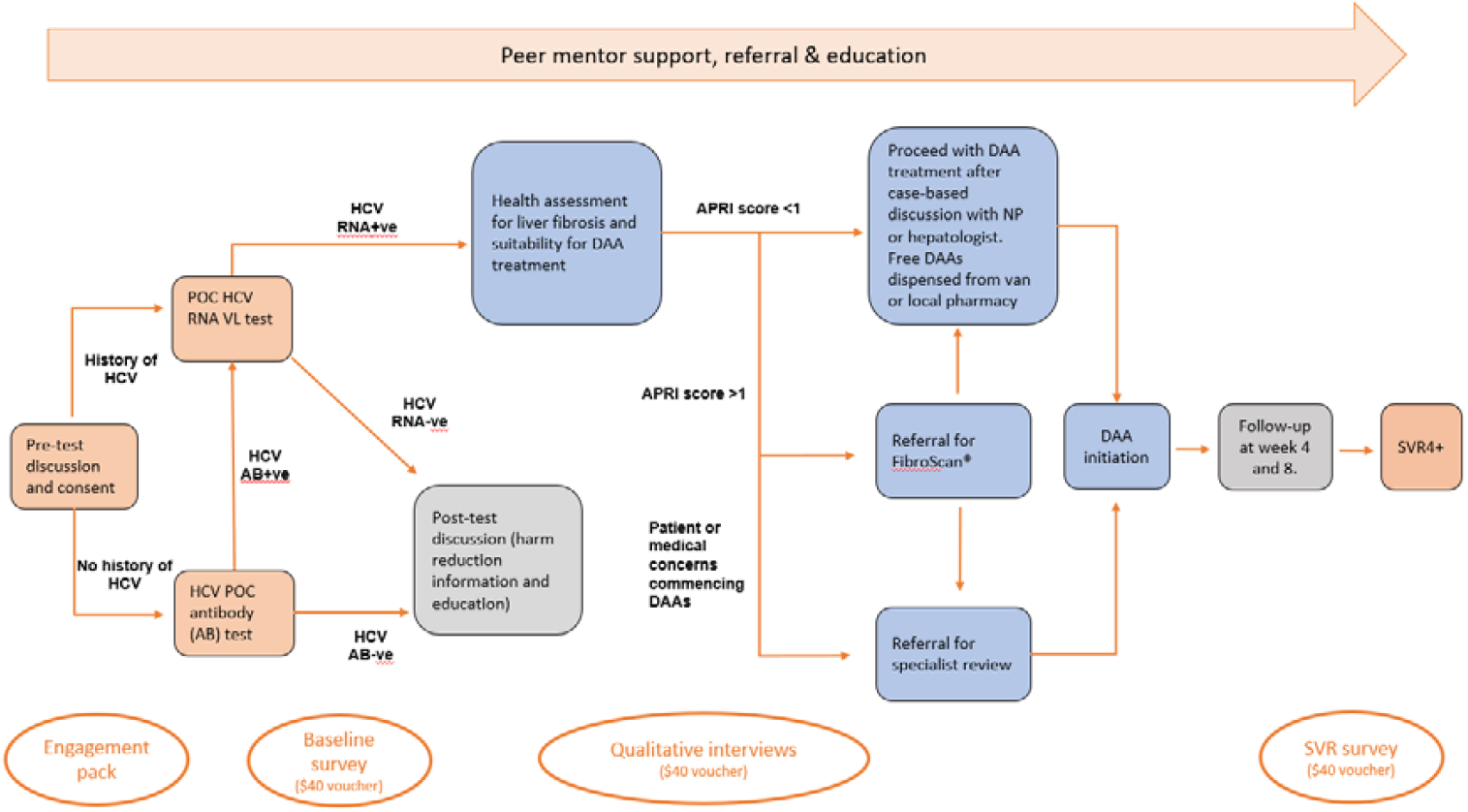
Participant pathway and timeline through C No More testing and treatment.

An authority prescription for DAA treatment will be provided to a pharmacy of the participants’ choice, a local partner pharmacy, or will be dispensed through the hospital pharmacy, and provided to participants through the van. The quantity of the DAA treatment course dispensed will depend on personal circumstances and participant preference, ranging from monthly supply to the full treatment course. Participants will be supported through dispensation and treatment by the study nurse and/or peer worker by phone, at commencement, at four-week intervals during treatment, and additionally as needed.

### Post-treatment assessment for sustained virological response

Participants who commence treatment will be invited for clinical assessment at least four weeks after completion of their treatment regimen. Participants will undertake point-of-care hepatitis C RNA testing with the GeneXpert® HCV VL Fingerstick assay to determinesustained virological response (SVR-4+). If participants cannot attend the mobile clinic, they will be mailed a pathology request for SVR4+ testing via a local pathology provider, with results to be provided to the study.

### Other study related procedures

#### Demographic and risk behaviours questionnaire

Participants will be asked to complete a voluntary researcher-administered short survey that collects data on participant demographics, health status, drug and alcohol use, social support, risk factors for hepatitis C infection, and previous hepatitis C testing and treatment. This survey was developed for this study (Supplementary 2). This survey will be offered at the time of consent and hepatitis C testing. If applicable, a post-treatment survey is offered at the post-treatment SVR assessment, focussing on recent risk factors and experience with the ‘C No More’ model of care. Participants who elect to complete the baseline and SVR surveys will be provided with a $40 Coles Myer voucher per survey as reimbursement for their time.

#### Qualitative appraisal

When consenting into the study, participants will be asked if they consent to be contacted at a future time to participate in further studies. Participants who opt-in to being contacted will be considered for further qualitative interviews. We will aim to conduct in-depth, semi-structured interviews with a subset of 20 participants who agree to them. We will use purposive sampling to ensure the inclusion of information-rich cases and a breadth of experiences, including balance across gender, criminal justice history and community corrections site. Interviews will be guided by a semi-structured schedule that focuses on participant experiences and perspectives of the ‘C No More’ mobile model of care, prior experiences with hepatitis C care, and prior experiences with traditional healthcare providers and settings. Participation in in-depth interviews will be voluntary, will include informed written consent, and participants will be offered $40 Coles Myer vouchers as a reimbursement for their time.

#### Harm reduction

Participants will be offered harm reduction advice and materials including naloxone and clean injecting equipment by the peer workers and the study nurse. The peer workers will source and maintain naloxone and injecting equipment stock.

### Outcomes

Model of care feasibility and effectiveness will be assessed using the following indicators:

1. The number of INSTI® or Abbott Bioline® HCV Antibody tests performed;
2. The proportion of individuals tested who are positive for HCV antibody;
3. The number of GeneXpert® HCV RNA tests performed;
4. The proportion of individuals tested who are positive for hepatitis C RNA;
5. The proportion of HCV RNA individuals who are commenced on DAA treatment;
6. The proportion of HCV RNA positive individuals who complete DAA treatment;
7. The proportion of individuals who achieve sustained virological response (SVR4+ (defined as HCV RNA negative at least 4 weeks post treatment).

Model of care acceptability will be assessed using the following indicators:

1. Survey responses describing the population accessing hepatitis C point-of-care testing at the mobile van, including sociodemographics, risk factors for hepatitis C, contact with the justice system including criminal justice history, drug use and protective behaviours, and any prior diagnosis and treatment of hepatitis C;
2. Qualitative evaluation interviews with individuals accessing the service.

### Study timeline

#### Sample size

We will aim to recruit 650 participants over 18 months. Based on international data (23), we estimate that 7% (n=45) will be HCV RNA positive and will be eligible to initiate treatment. A sample of 650 will be sufficient to demonstrate superiority of the proportion of HCV RNA positive individuals who initiate treatment in the intervention model relative to an Australian primary care sentinel surveillance estimate (51% treatment initiation), presuming 34% margin with 90% power at the 5% significance level. The primary care estimate of treatment initiation will be sourced from The Australian Collaboration for Coordinated Enhanced Sentinel Surveillance of Sexually Transmissible Infections and Blood Borne Viruses (ACCESS) database. The national estimate for the hepatitis C cascade of care in primary care clinics has been published (5, 27), however we will use an unpublished subset of data, restricted to Victorian primary care clinics for comparison.

#### Recruitment

Individuals in the vicinity of corrections offices, including those attending community corrections offices, will be opportunistically approached by the peer worker and invited to undertake hepatitis C testing. If an individual indicates interest in being tested, they will be invited into the van where the study protocol and aims will be discussed with them. If the individual wishes to take part, they will be provided with a Participant Informed Consent Form (Supplementary 3) and if they choose to participate, signed informed consent will be obtained. Participants will be provided with an engagement pack to encourage participation, which contains toiletries and snacks.

Information sessions incorporating education on hepatitis C transmission, infection sequelae, epidemiology, prevention, testing, treatment and the mobile model of care will be provided to community corrections staff, who will be encouraged to promote the service to their clients. Promotional materials will be provided to the corrections offices for display in waiting areas and consultation rooms.

#### Participant retention

The study nurse will confirm that dispensing has occurred with the dispensing pharmacy. Study staff will follow-up with participants on treatment by phone at four-weekly intervals to check the participant is still taking their treatment and additional support will be offered for participants who struggle to comply with treatment guidelines. Study staff will make repeated attempts to follow-up participants who are unable to be contacted. At enrolment, participants who are undertaking an HCV RNA test or commencing treatment, will be asked to provide secondary contact information, such as a friend, family member, or support worker such as social worker or corrections worker, who they would be happy for study staff to contact in the event they are unable to be directly contacted.

#### Staff training and quality assurance

All study staff that interact with participants will be trained in data collection and obtaining informed consent. Staff will be trained in performing both INSTI®/Bioline® hepatitis C antibody and GeneXpert® hepatitis C RNA testing. Standard quality assurance is being performed for the GeneXpert HCV RNA® machine, in line with the National Point of Care Testing program requirements.

#### Data collection methods

All data will be collected and recorded electronically using REDCap® electronic data capture tools hosted by Burnet Institute. REDCap® is a secure, web-based software data collection platform (45, 46). Minimal contact details and clinical data will be collected for participants undertaking only an HCV antibody test, prior to performing the test. For participants who undertake GeneXpert® HCV RNA testing, further clinical and contact data will be collected to enable follow-up, if needed. Data from routine clinical care will be recorded on paper or electronic data collection record forms at each visit and entered by study personnel into REDCap®.

Demographic, health, social, and behavioural information will be collected for participants consenting to undertake the baseline questionnaire and entered into REDCap®.

#### Data management

Identifiable and contact information will be stored separately to clinical, demographic, and risk-factor information. Clinical, demographic, and risk-factor information will be anonymised with an automatically generated study ID and stored in a separate REDCap® database.

#### Data linkage

Participants’ data collected in the baseline questionnaire and clinical data tools will be prospectively linked with the following administrative data sources where participants have consented to this:

1. Medicare (access to Medicare-subsidised health services);
2. Pharmaceutical Benefits Scheme (government-subsidised medication prescriptions); and
3. Corrections data (periods of Corrections supervision, community-based supervision and incarceration)

### Statistical analyses

Feasibility and effectiveness of the model of care will be determined by using clinical data to determine the number and proportion of individuals consenting to be tested, the number of HCV antibody and RNA tests performed, the number and proportion of individuals who are positive for HCV RNA, the number and proportion of HCV RNA positive individuals who initiate DAA treatment, the number and proportion of individuals who complete treatment, and the number and proportions of individuals that achieve SVR4+.

Descriptive data on the study population will be presented using numbers (proportion) for categorical data, and median (interquartile range) for continuous data. Categorical variables will be assessed using chi-squared test, or Fisher’s exact test for small populations. Continuous variables will be applied to t-tests and Mann-Whitney/Kruskal-Wallis tests. Data will be analysed using Stata v18.0 (StataCorp LLC).

Model of care acceptability will be determined through in-depth qualitative evaluation interviews with a range of individuals accessing the service. Qualitative data will be coded in NVivo 14 (Lumivero) and analysed thematically in according to a healthcare acceptability framework.

## Data Availability

The participants of this study did not give written consent for their data to be shared publicly, so due to the sensitive nature of the research supporting data is not available.

## Declarations

### Ethics approval

This study has been approved by the St Vincent’s Hospital Melbourne Human Research Ethics Committee (ref:025/023).

### Informed consent

People eligible to participate in the study will be formally consented inside the van. Trained study staff will provide a detailed overview of the contents of the Participant Informed Consent Form (PICF), and a detailed overview of the study.

Potential participants will be reminded that participation is entirely voluntary, and any questions that arise will be answered by study staff. Potential participants will be asked to provide written consent via the PICF, which will be recorded and stored online via REDCap. Participants will be asked to consent to have their survey data linked to Medicare and Pharmaceutical Benefits Scheme data, to monitor hepatitis C testing, positivity, and treatment via other providers to determine if treatment was accessed from an alternate health service in the event of loss-to-follow-up, and to observe new primary and reinfections. Participants will also be asked to consent to linkage to Corrections Victoria data, to monitor future periods of incarceration and determine if testing and/or treatment was accessed while incarcerated.

### Consent for publication

Not applicable.

### Availability of data and materials

Not applicable.

### Protocol amendments

Any modifications to the protocol that may impact on the conduct of the study, potential benefit to the patient, or may affect patient safety will require a formal amendment to the protocol and will be approved by St Vincent’s Hospital Melbourne Human Research Ethics Committee.

### Study withdrawal

All study participants may exercise their right to withdraw from the study. Participants who wish to withdraw will be asked to complete a withdrawal of consent form and will be asked to discuss their reasons for withdrawal, and use of their previously collected personal and health information data.

### Privacy and confidentiality

Participants will be assigned a unique, automatically generated study identification number when enrolled into the study, with all contact details and potentially identifying data stored separately to clinical or questionnaire data. All data will be stored in a secure, password-protected database, with access limited to personnel directly involved in the conduct of the study. Any paper files relating to the study will be stored in a locked filing cabinet in the study co-ordinator’s office. No identifying information will be included in any report, presentation, or publication relating to the findings of the study.

Clinical assessments and participant surveys will be conducted individually, and away from other participants where necessary, to protect individuals’ privacy and confidentiality during clinical assessments and interviews.

### Dissemination

As per standard care, all clinical investigation results will be reported confidentially to participants. The results of this study will be published in peer-reviewed journals and/or presented at scientific meetings. Results may form part of PhD or other student theses. All quantitative data will be presented in non-identifiable, aggregate format. Qualitative data will be presented in a non-identifiable format.

### Author contributions

SG: Project administration, writing-original draft.

JAH: Conceptualisation, Investigation, Supervision, Writing-review and editing.

BR: Investigation, Writing-review and editing.

TP: Conceptualisation, Writing-review and editing.

ACr: Investigation, Writing-review and editing.

JD: Resources, conceptualisation. AC: Investigation.

MB: Investigation.

SW: Conceptualisation, writing-review and editing.

MH: Writing-review and editing.

AJT: Conceptualisation, Investigation, Supervision, Writing-review and editing.

MS: Conceptualisation, Supervision, Writing-review and editing.

RJW: Conceptualisation, Project administration, Supervision, Writing-review and editing.

## Acknowledgements

The clinical implementation of this study is funded by Gilead sciences Pty Ltd via an independent medical grant. The evaluation of this study is supported by St Vincent’s Hospital and National Health and Medical Research Council. The study also receives in-kind support from the National Point-of-care Testing Program and Kirby Institute.

## List of abbreviations

APRI: Aspartate aminotransferase to Platelet Ratio Index
DAA: direct-acting antiviral
DDI: drug-drug interactions
HCV: Hepatitis C Virus
SVR-4+: sustained virological response 4+ weeks after treatment completion
WHO: World Health Organization

## Appendices

Supplementary 1. SPIRIT statement

Supplementary 2. Questionnaire

Supplementary 3. Informed consent materials

Supplementary 4. Biological specimens

